# Effectiveness of inactive COVID-19 vaccines against severe illness in B.1.617.2 (Delta) variant-infected patients in Jiangsu, China

**DOI:** 10.1101/2021.09.02.21263010

**Authors:** Zhiliang Hu, Bilin Tao, Zhongqi Li, Yan Song, Changhua Yi, Junwei Li, Meng Zhu, Yongxiang Yi, Peng Huang, Jianming Wang

## Abstract

The SARS-CoV-2 B.1.617.2 (Delta) variant has caused a new surge in the number of COVID-19 cases. The effectiveness of vaccines against this variant is not fully understood. Using data from a recent large-scale outbreak of COVID-19 in China, we conducted a real-world study to explore the effect of inactivated vaccine immunization on the course of disease in patients infected with Delta variants. We recruited 476 confirmed cases over the age of 18, of which 42 were severe. After adjusting for age, gender, and comorbidities, patients who received two doses of inactivated vaccine (fully vaccinated) had an 88% reduced risk in progressing to the severe stage (adjusted OR: 0.12, 95% CI: 0.02- 0.45). However, this protective effect was not observed in patients who only received only one dose of the vaccine(adjusted OR: 1.11, 95% CI: 0.51- 2.36). The full immunization offered 100% protection from a severe illness among women. The effect of the vaccine was potentially affected by underlying medical conditions (OR: 0.26, 95% CI: 0.03-1.23). This is the largest real-world study confirming the effectiveness of inactive COVID-19 vaccines against severe illness in Delta variant-infected patients in Jiangsu, China.

## Introduction

According to estimation by World Health Organization (WHO), as of August 24, 2021, the global cumulative number of confirmed coronavirus disease 2019 (COVID-19) cases has risen to over 210 million, and more than 4.4 million people have died from it[1]. There is no doubt that vaccination is a vital measure to contain the global COVID-19 pandemic. Different COVID-19 vaccines, including inactivated, adenovirus vector, and mRNA vaccines, have been authorized or in the laboratory development and clinical utility evaluation stage[2, 3, 4, 5]. As of August 23, 2021, over 4.6 billion doses of the COVID-19 vaccines have been administered globally[1]. These vaccines can effectively induce immune responses against severe acute respiratory syndrome coronavirus 2 (SARS-CoV-2) infection[6, 7, 8]. Clinical trials outside China demonstrated that vaccine efficacy for preventing symptomatic COVID-19 ranged from 65.9% to 83%, and severe illness or ICU admission ranged from 90% to 100%[9, 10, 11]. In China, the masses are generally vaccinated with inactivated vaccines. Accumulated evidence suggests that inactivated COVID-19 vaccine could efficiently, although not wholly, protect from SARS-CoV-2 infection and, importantly, prevent severe illness progression. However, it is difficult to confirm in mainland China because there was no large-scale local outbreak after the first epidemic wave in 2020. Moreover, the protective effect of the inactivated vaccine on the pathogenesis of SARS-CoV-2 mutant strains is not clear.

On July 20, 2021, nine domestic COVID-19 cases were identified through a regular screening in Nanjing Lukou international airport, China[12]. The epidemic gathered at the airport spread rapidly to the surrounding areas, leading to outbreaks in Nanjing, Yangzhou, and Zhangjiajie. Genome sequencing confirmed that the etiologic agent was the SARS-CoV-2 B.1.617.2 (Delta) variant, which was first identified in Maharashtra, India, in late 2020 and now has spread globally[13]. Compared with the original type of SARS-CoV-2, the Delta variant has a significantly increased virulence and transmission ability[14, 15, 16]. Furthermore, the protective effect of vaccines against Delta variant infection is found to be weakened in many studies[17, 18, 19, 20, 21].

In the case of more than 200 million doses of COVID-19 vaccine have been administered in China, whether the widely used inactivated vaccine is still effective for Delta variants is a question worthy of discussion. Thus, we performed a real-world study using patients' clinical and epidemiological data in a designated hospital in Nanjing. They were all linked to the outbreak of COVID-19 in Nanjing Lukou international airport. Our study aimed to describe to what extent the inactivated vaccine could prevent COVID-19 from progressing to severe illness in patients infected with the SARS-CoV-2 Delta variant.

## Methods

### Study design and population

We recruited 476 confirmed COVID-19 cases treated in the isolation wards of Nanjing Public Health Medical Center from June to July. The inclusive criteria were patients: 1) aged over 18 years, 2) confirmed by SARS-CoV-2 nucleic acid PCR test, 3) linked to the recent outbreak of COVID-19 originated in Nanjing Lukou international airport, 4) infected with Delta variant. The Nanjing Public Health Medical Center is the only designated hospital that provides medical services for COVID-19 patients in Nanjing. Of the 476 patients recruited in this study, 189 lived in Nanjing, 273 lived in Yangzhou, 12 lived in Huaian, and two lived in Suqian. We collected data of each patient, including demographic characteristics, medical history, vaccine status, comorbidities, clinical features, laboratory tests, treatments, and outcomes. The onset date was defined as when symptoms first appeared, or asymptomatic patients were detected for the first time with SARS-CoV-2 nucleic acid positive. The diagnosis of severe illness was based on the “Guideline of COVID-19 Diagnosis and Treatment (trial version 8)” issued by the National Health Council of China. This study was approved by the ethics committee of Nanjing Public Health Medical Center. Written informed consent was waived by the Ethics Commission.

### Vaccination status

The information regarding the time of vaccination and the type of the vaccines were obtained from the electronic health information system. The time interval between the last dose of vaccination and the onset of disease was calculated. Because two weeks after the second dose were needed to develop protective immune responses against SARS-CoV-2 infection, an effective vaccine shot was considered only when the time interval between the second shot and disease onset was at least 14 days. We categorized patients into three groups: unvaccinated, partially vaccinated, and fully vaccinated according to the immunization history. Patients would also be considered unvaccinated if they had received one dose but the time interval between the first shot and illness onset was less than 14 days. Likewise, patients, who had received two vaccine shots, however, the time interval between the second shot and illness onset was less than 14 days, would be considered partially vaccinated[22].

### Outcomes

The primary outcome of interest was the progression to severe illness in patients infected with the Delta variant. As defined by the “Guideline of COVID-19 Diagnosis and Treatment (trial version 8)” in China, severe illness of COVID-19 for adult patients should meet one of the following criteria: 1) respiratory rate ≥30 breaths/min, 2) at rest, the oxygen saturation of fingers during air inhalation is ≤93%, 3) arterial partial pressure of oxygen (PaO_2_) / oxygen uptake concentration (FiO_2_) ≤300 mmHg, 4) clinical symptoms were aggravated, and the pulmonary imaging showed that the lesion progressed more than 50% within 24-48 hours. Patients with critical COVID-19 were those who had developed respiratory failure requiring mechanical ventilation or had evidence of shock or other organ dysfunctions that needed transferring to the intensive care unit (ICU)[23]. The most severe condition of the patients during the hospitalization was recorded. In this study, we analyzed severe and critical cases together.

### Covariates

Covariates that have been confirmed to or possibly have a role in disease progression were considered, including age, gender, comorbidities, vaccination status, baseline SARS-CoV-2 viral load, and therapies (corticosteroids, intravenous immunoglobulin, and aerosol interferon-a). Age was categorized into two groups: 18-59 years and >=60 years. Clinical parameters such as blood lymphocyte counts, C-reactive protein(CRP), Interleukin-6 (IL-6), D-dimer, lactate dehydrogenase (LDH), and pulmonary involvement were more appropriate to an index of disease severity rather than the risk factors, thereby were not included in the multivariable regression analysis. All cases involved in this study were vaccinated with the inactivated vaccine.

### Statistical analysis

Categorized variables were expressed as frequency, and continuous variables were described as the median (interquartile range, IQR). Comparison between groups was made using the Chi-Square test, Mann-Whitney U-test, or Fisher’s exact test, as appropriate. Factors related to severe illness were analyzed by univariate and multivariate regression analysis, and the relationship was expressed with odds ratio (OR) and 95% confidence interval (95% CI). We also performed a subgroup analysis by stratifying age, gender, and underlying medical conditions. The significance level was set at 0.05. All analyses were performed using R software for Windows version 4.0.5 (https://www.r-project.org/).

## Results

### Demographics and clinical characteristics

A total of 476 hospitalized patients were included in the analysis, of which 42 (8.82%) were severe cases. Over 60% of patients were female. The median age of patients with severe and non-severe illnesses was 69.50 (IQR: 59.00-75.00) years and 51.00 (IQR: 40.00-66.00) years. Patients with severe illness tended to had more underlying medical conditions (50.00% vs. 30.02%), including hypertension (35.71% vs. 21.71%), diabetes (19.05% vs. 8.08%), heart disease (11.90% vs. 3.70%) and COPD (4.76% vs. 0.23%). No significant difference was observed between the two groups in gender and history of cancer, asthma, or autoimmune diseases. Time from disease onset to hospitalization was similar in patients with severe or non-severe illness, with a median time of 3 days. In patients without severe innless, there were 157 (36.18%) unvaccinated, 92 (21.20%) partially vaccinated, and 185 (42.63%) fully vaccinated. In contrast, in patients with severe illness, the proportion of unvaccinated, partially vaccinated, and fully vaccinated was 64.29%, 0.95%, and 4.76%, respectively. (Table 1).

**Table 1.** Baseline characteristics and clinical features of 476 Delta variant-infected patients with or without severe illness.

|  | Non-severe illness<br>n=434, n (%) or<br>median (IQR) | Severe illness<br>n=42, n (%) or median<br>(IQR) | P |
| --- | --- | --- | --- |
| **Demographics and clinical characteristics** |  |  |  |
| Gender |  |  | 0.321 |
| Female | 267 (61.52) | 22 (52.38) |  |
| Male | 167 (38.48) | 20 (47.62) |  |
| Age group, years | 51.00 (40.00, 66.00) | 69.50 (59.00-75.00) | <0.001 |
| 18-59 | 286 (65.90) | 12 (28.57) | <0.001 |
| ≥60 | 148 (34.10) | 30 (71.42) |  |
| Having underlying medical conditions | 130 (30.02) | 21 (50.00) | 0.013 |
| Hypertension | 94 (21.71) | 15 (35.71) | 0.062 |
| Diabetes | 35 (8.08) | 8 (19.05) | 0.037 |
| Heart disease | 16 (3.70) | 5 (11.90) | 0.038 |
| Carcinoma history | 9 (2.08) | 2 (4.76) | 0.253 |
| COPD | 1 (0.23) | 2 (4.76) | 0.022 |
| Asthma | 8 (1.85) | 1 (2.38) | 0.569 |
| Autoimmune diseases | 4 (0.93) | 1 (2.38) | 0.372 |
| Vaccination Status |  |  | <0.001 |
| Unvaccinated | 157 (36.18) | 27 (64.29) |  |
| Partially vaccinated | 92 (21.20) | 13 (30.95) |  |
| Fully vaccinated | 185 (42.63) | 2 (4.76) |  |
| Time from illness onset to<br>hospitalization (days) | 3.00 (1.00, 5.00) | 3.00 (2.00,5.00) | 0.152 |
| **Clinical features** |  |  |  |
| Fever | 138 (31.87) | 26 (61.90) | <0.001 |
| Cough | 221 (51.04) | 23 (54.76) | 0.765 |
| Sputum production | 84 (19.35) | 8 (19.05) | 1.000 |
| Shortness of breath | 15 (3.46) | 8 (19.05) | <0.001 |
| Nausea or vomiting | 9 (2.08) | 2 (4.76) | 0.253 |
| Abdominal pain or diarrhea | 25 (5.76) | 3 (7.14) | 0.728 |
| Loss of smell or taste | 18 (4.15) | 1 (2.38) | 1.000 |
| Myalgia | 11 (2.53) | 3 (7.14) | 0.117 |
| Stuffy nose or runny nose | 64 (14.75) | 3 (7.14) | 0.245 |
| Headache or dizziness | 30 (6.91) | 5 (11.90) | 0.382 |
| Fatigue | 91 (20.97) | 8 (19.05) | 0.925 |
| Pharyngeal discomfort | 97 (22.35) | 6 (14.29) | 0.310 |
| **Laboratory findings** |  |  |  |
| C-reactive protein , mg/L | 4.96 (1.89, 13.99) | 25.94 (11.22-36.50) | <0.001 |
| >10 | 288 (66.36) | 10 (23.81) | <0.001 |
| Procalcitonin, ng/mL | 0.04 (0.02, 0.06) | 0.09 (0.06-0.14) | <0.001 |
| <0.1 | 385 (89.33) | 26 (61.90) |  |
| 0.1- | 43 (9.98) | 12 (28.57) |  |
| 0.25- | 2 (0.46) | 2 (4.76) |  |
| 0.5- | 1 (0.23) | 2 (4.76) |  |
| Interleukin-6, pg/mL | 10.25 (3.48, 20.52) | 37.67 (22.00-72.61) | <0.001 |
| >6.6 | 159 (36.89) | 1 (2.38) | <0.001 |
| White blood cell count, × 10<sup>9</sup>/L | 4.81 (3.88, 5.93) | 4.67 (3.99,5.96) | 0.779 |
| <4 | 128 (29.49) | 11 (26.19) | 0.869 |
| 4 | 298 (68.66) | 31 (73.81) |  |
| 10- | 8 (1.84) | 0 (0.00) |  |
| Neutrophil count, × 10<sup>9</sup>/L | 3.04 (2.18, 3.93) | 3.22 (2.37,4.17) | 0.423 |
| Lymphocyte count, $\times 10^9/L$ | 1.16 (0.86, 1.56) | 1.04 (0.88, 1.27) | 0.063 |
| <0.8 | 78 (17.97) | 8 (19.05) | 1.000 |
| Hemoglobin, g/L | 131.00 (122.00, 145.00) | 132.00 (123.25, 138.00) | 0.617 |
| Platelet count, $\times 10^9/L$ | 157.00 (123.25, 193.75) | 117.50 (92.50, 144.75) | <0.001 |
| <100 | 54 (12.44) | 14 (33.33) | 0.001 |
| Albumin, g/L | 42.60 (40.30, 45.20) | 40.10 (37.30, 42.18) | <0.001 |
| Total bilirubin, $\mu\text{mol/L}$ | 9.80 (7.70, 12.40) | 10.10 (8.33, 13.05) | 0.532 |
| Aspartate aminotransferase, IU/L | 17.05 (13.83, 23.28) | 25.15 (19.72, 37.70) | <0.001 |
| >40 | 35 (8.06) | 9 (21.43) | 0.010 |
| Alanine aminotransferase, U/L | 17.85 (12.33, 28.25) | 23.80 (14.10, 32.05) | 0.038 |
| >40 | 57 (13.13) | 7 (16.67) | 0.686 |
| Urea nitrogen, mmol/L | 4.44 (3.64, 5.29) | 5.10 (3.84, 6.45) | 0.012 |
| Creatinine, $\mu\text{mol/L}$ | 60.25 (51.80, 74.80) | 71.20 (59.50, 81.68) | 0.001 |
| >104 | 11 (2.53) | 3 (7.14) | 0.117 |
| eGFR, ml/min | 111.38 (101.28, 121.21) | 92.85 (83.95, 107.70) | 0.001 |
| <90 | 31 (10.10) | 7 (43.75) | <0.001 |
| Creatine kinase, U/L | 67.50 (47.00, 106.00) | 133.00 (64.25, 209.75) | <0.001 |
| >174 | 47 (10.93) | 15 (35.71) | <0.001 |
| CK-MB, ng/ml | 0.50 (0.30, 0.90) | 0.60 (0.40, 0.90) | 0.183 |
| >3.1 | 11 (2.94) | 1 (2.38) | 1.000 |
| Myoglobin, ng/mL | 33.40 (21.25, 58.10) | 66.50 (38.10, 107.20) | <0.001 |
| >106 | 37 (9.97) | 11 (26.83) | 0.003 |
| Troponin I, pg/ml | 3.00 (0.72, 6.65) | 10.10 (4.50, 18.60) | <0.001 |
| >15.6 | 29 (7.92) | 14 (34.15) | <0.001 |
| Lactate dehydrogenase, U/L | 235.00 (204.00, 274.00) | 312.50 (240.00, 378.75) | <0.001 |
| >245 | 188 (43.32) | 31 (73.81) | <0.001 |
| Prothrombin time, s | 11.10 (10.80, 11.60) | 11.50 (11.10, 11.80) | 0.003 |
| D-dimer, $\mu\text{g/L}$ | 0.39 (0.25, 0.64) | 0.46 (0.32, 0.88) | 0.018 |
| >0.55 | 132 (30.41) | 18 (42.86) | 0.138 |
| FDPs, $\mu\text{g/ml}$ | 2.70 (1.77, 5.33) | 3.55 (2.70, 5.35) | 0.120 |
| **Microbiological and serological data** |  |  |  |
| Viral load (Ct value) |  |  |  |
| ORF1ab gene | 23.00 (19.00, 28.00) | 23.00 (21.00, 26.75) | 0.633 |
| N gene | 20.00 (16.00, 25.00) | 20.00 (16.25, 23.00) | 0.850 |
| Antibody levels, S/CO |  |  |  |
| SARS-COV-2 IgM antibody | 0.24 (0.06, 1.19) | 0.08 (0.05, 0.62) | 0.049 |
| SARS-COV-2 IgG antibody | 1.57 (0.15, 17.08) | 0.12 (0.05, 0.31) | <0.001 |
| **Treatments** |  |  |  |
| Corticosteroids | 0 (0.00) | 1 (2.38) | 0.088 |
| Intravenous immunoglobulin | 1 (0.23) | 1 (2.38) | 0.169 |
| Thymosin | 52 (11.98) | 6 (14.29) | 0.850 |
| Arbidol | 40 (9.22) | 2 (4.76) | 0.566 |
| Aerosol interferon-alpha | 38 (8.76) | 1 (2.38) | 0.235 |

1. Data were expressed as median (interquartile range), or n (%). As appropriate, a comparison between groups was made using Mann-Whitney U-test, Chi-Square test, or Fisher's exact test.
2. COPD, chronic obstructive pulmonary disease; CK-MB, creatine kinase-myocardial band; eGFR, estimated glomerular filtration rate; FDPs, fibrinogen degradation products; Ct, cycle threshold; SARS-COV-2, severe acute respiratory syndrome coronavirus 2.
3. the Ct value from quantitative reverse transcription polymerase chain reaction was used to represent the viral load of SARS-COV-2 in the upper respiratory tract.

### Signs and symptoms

Initial symptoms were fever, dry cough, sputum production, chest tightness or shortness of breath, nausea or vomiting, abdominal pain or diarrhea, loss of smell or taste, myalgia, stuffy or runny nose, headache or dizziness, fatigue, and pharyngeal discomfort. Patients with severe illness had a higher proportion of fever (61.90% vs. 31.87%) and shortness of breath (19.05% vs. 3.46%) than those without severe illness. Besides, the patients with severe illness were commonly cough, sputum production, nausea or vomiting, abdominal pain or diarrhea, myalgia, headache, or dizziness, but the difference was not statistically significant (Table 1).

### Laboratory parameters, microbiological and serological data

As shown in Table 1, patients who progressed to severe illness had relatively higher levels of baseline CRP, procalcitonin, IL-6, aspartate aminotransferase, alanine aminotransferase, urea nitrogen, creatinine, creatine kinase, myoglobin, troponin I, LDH, prothrombin time, and D-dimer. Baseline levels of the white blood cell count, lymphocyte count, neutrophil count, hemoglobin, total bilirubin, CK-MB, and fibrinogen degradation products were similar between the two groups. Patients without severe illness had a higher IgG and IgM antibody to SARS-CoV-2. There were no significant differences in viral load threshold cycle value (Ct) detecting ORF1ab and O gene.

### Estimate the efficacy of inactivated SARS-CoV-2 vaccine

Severe illness occurred in 14.7% (27/184), 12.4% (13/105), and 1.1% (2/187) of the unvaccinated, partially vaccinated) and fully vaccinated patients, respectively. As pre-defined in the methods, patients who had received one dose of vaccine and had acquired Delta variant infection within 14 days were deemed unvaccinated. This 14-days elapsed time was also applicable to the second dose vaccination. There was no significant difference in the proportion of severe illness between patients who did not receive any COVID-19 vaccine and patients having one dose within 14 days (15.6% vs. 12.2%, P =0.75), or between patients who did not receive any COVID-19 vaccine and patients having the 2nd dose within 14 days (12.8% vs. 11.1 %, P =1). Therefore, our estimation of the effectiveness of vaccines would not be significantly biased by the definition of vaccination status.

Compared with the unvaccinated group, those fully vaccinated ones had a significantly decreased risk of severe illness (OR: 0.06, 95% CI: 0.01-0.21, P<0.001; risk reduction: 94%, 95% CI: 79%-99%). The risk of severe illness was also decreased for the partially vaccinated person but not significant (OR: 0.822, 95% CI: 0.393-1.643).

After adjusting for the potential confounders, such as gender, age, and underlying medical conditions, the protective effect of full vaccination remained significant (adjusted OR: 0.12, 95% CI: 0.02- 0.45, P=0.006; adjusted risk reduction: 88%, 95%CI: 55%-98%). No significant effect was found for partial vaccination (adjusted OR: 1.11, 95% CI: 0.51- 2.36, P=0.783) (Table 2).

**Table 2.** Effectiveness of inactivated COVID-19 vaccines against severe illness in patients infected with Delta variant.

|  | Unvaccinated<br>(Ref.) | Partially vaccinated |  |  |  |  | Fully vaccinated |  |  |  |  |
| --- | --- | --- | --- | --- | --- | --- | --- | --- | --- | --- | --- |
|  | N | N | OR (95%CI) | P | OR<sub>adj</sub> (95%CI) | P<sub>adj</sub> | N | OR (95%CI) | P | OR<sub>adj</sub> (95%CI) | P<sub>adj</sub> |
| **All patients** |  |  |  |  |  |  |  |  |  |  |  |
| Non-Severe illness | 157 | 92 |  |  |  |  | 185 |  |  |  |  |
| Severe illness | 27 | 13 | 0.82<br>(0.39-1.64) | 0.59 | 1.11<br>(0.51-2.36) | 0.78 | 2 | 0.06<br>(0.01-0.21) | < 0.001 | 0.11<br>(0.02-0.45) | 0.006 |
| **With underlying medical conditions** |  |  |  |  |  |  |  |  |  |  |  |
| Non-Severe illness | 68 | 31 |  |  |  |  | 31 |  |  |  |  |
| Severe illness | 15 | 4 | 0.59<br>(0.16-1.77) | 0.374 | 0.544<br>(0.14-1.77) | 0.34 | 2 | 0.29<br>(0.04-1.12) | 0.12 | 0.26<br>(0.03-1.23) | 0.121 |
| **Without underlying medical conditions** |  |  |  |  |  |  |  |  |  |  |  |
| Non-Severe illness | 89 | 60 |  |  |  |  | 154 |  |  |  |  |
| Severe illness | 12 | 9 | 1.11<br>(0.43-2.79) | 0.821 | 1.83<br>(0.65-5.10) | 0.247 | 0 | 0 | - | 0 | - |
| **18-59 years** |  |  |  |  |  |  |  |  |  |  |  |
| Non-Severe illness | 51 | 64 |  |  |  |  | 171 |  |  |  |  |
| Severe illness | 5 | 5 | 0.80<br>(0.21-3.00) | 0.731 | 0.67<br>(0.17-2.66) | 0.562 | 2 | 0.12<br>(0.02-0.57) | 0.013 | 0.12<br>(0.02-0.61) | 0.016 |
| **≥60 years** |  |  |  |  |  |  |  |  |  |  |  |
| Non-Severe illness | 106 | 28 |  |  |  |  | 14 |  |  |  |  |
| Severe illness | 22 | 8 | 1.38<br>(0.53-3.33) | 0.491 | 1.37<br>(0.52-3.31) | 0.502 | 0 | 0 | - | 0 | - |
| **Male** |  |  |  |  |  |  |  |  |  |  |  |
| Non-Severe illness | 57 | 43 |  |  |  |  | 67 |  |  |  |  |
| Severe illness | 12 | 6 | 0.66<br>(0.22-1.85) | 0.446 | 0.80<br>(0.24-2.48) | 0.702 | 2 | 0.14<br>(0.02-0.55) | 0.013 | 0.19<br>(0.03-0.86) | 0.049 |
| **Female** |  |  |  |  |  |  |  |  |  |  |  |
| Non-Severe illness | 100 | 49 |  |  |  |  | 118 |  |  |  |  |
| Severe illness | 15 | 7 | 0.95<br>(0.35-2.42) | 0.921 | 1.37<br>(0.47-3.74) | 0.545 | 0 | 0 | - | 0 | - |

OR, odds ratio; CI, confidence interval; OR<sub>adj</sub>, adjusted odds ratio

### Subgroup analysis

The risk of progressing to severe illness reached 0 in fully vaccinated persons without underlying medical conditions, age≥60 years or female. Only 14 elderly patients were fully vaccinated, so that the protection may be overestimated in this subgroup. The protective effect of severe illness remained significant for 18-59 years fully vaccinated persons (adjusted OR: 0.12, 95% CI: 0.02-0.61, P: 0.016; risk reduction: 88%, 95% CI: 39%-98%) and male fully vaccinated persons (adjusted OR: 0.19, 95% CI: 0.03-0.86, P=0.049; risk reduction: 81%, 95% CI: 14%-97%). The effect of the vaccine was potentially affected by underlying medical conditions, resulting in reduced protective effect of full vaccination (OR: 0.26, 95% CI: 0.03-1.23). Partial vaccination had no significant protective effect on severe illness in all subgroups (P >0.05) (Table 2).

## Discussion

Mutations of the SARS-CoV-2 have attracted significant public attention, with variants of concerns leading to increased transmissibility, impaired immune protection by the vaccine, more severe disease, or compromised diagnostic capacity[24]. The Delta variant, which was first identified in India, is more transmittable than other lineages of SARS-CoV-2 and now is becoming the major strain causing the COVID-19 pandemic [25, 26, 27]. Vaccine breakthrough caused by the Delta variant has been increasingly reported, even in the massively vaccinated regions [27, 28]. Therefore, the concern has been raised regarding whether herd immunity in China established extensively by inactive vaccines could protect against the Delta variant.

In late May, the first attack by this new virus occurred in Guangzhou, China, with about 160 cases involved[29]. A real-world study on 74 patients and 292 negative controls calculated that the overall effect for two-dose vaccination was 59.0% against SARS-CoV-2 infection and 100% against severe illness[22]. Our present study, focusing on 476 hospitalized patients, demonstrated that the risk of progression to severe illness had substantially decreased in fully vaccinated patients. After adjusting for age, sex, and underlying medical conditions, the risk reduction remained significant, with an 88% risk reduction. This is the largest real-world study to confirm the effectiveness of inactivated vaccines in preventing severe illness caused by Delta variants in China.

It is well known that underlying comorbidities and old age are risk factors for severe illness in SARS-CoV-2 infected patients[30]. This is consistent with findings from our study. Severe illness did not occur in fully vaccinated patients without underlying medical conditions (100% protection). Both of the two fully vaccinated patients who developed severe illness had underlying diseases. Interestingly, 100% protection was also found in elderly patients that had been fully vaccinated. Since only 14 elderly patients were fully vaccinated, the protective effect of inactive vaccines might be overestimated in this study. Fully vaccinated women were 100% prevented from progressing to severe illness, while fully vaccinated men had only 81% reduced risk. Whether sex disparities exist in COVID-19 vaccine efficacy needs to be further explored.

Although an entire course of vaccination could efficiently protect the COVID-19 patients from progressing to severe illness, the protective effect could not be identified in partially vaccinated ones. This may be due to a relatively high viral burden and a decreased immune protection in Delta variant-infected patients [19, 20, 29]. The baseline viral load in this study, as represented by Ct value of the real-time quantitative reverse transcription polymerase chain reaction (RT-PCR) was 20 (IQR: 16-25), which is much higher than that (median: 30; IQR: 25-34) in our previous data during the first outbreak of COVID-19 in 2020[31]. In the context of Delta variant infection, a relatively higher immunity may be necessary, which generally would be achieved after a full vaccination[8].

There are some limitations to our study. First, we confirmed the protective effect of inactivated vaccines from progressing to severe illness, but we could not estimate the vaccine efficacy against Delta variant infection because all participants were confirmed COVID-19 cases. Second, since individuals who have been protected from infection would not develop a severe illness related to COVID-19, the effectiveness of inactive vaccines against severe illness in our study based on infected cases would be, to some extent, an underestimation of that based on the whole population.

In conclusion, we found a full course immunization with inactivated vaccines could effectively protect against severe illness caused by the Delta variant in China. The protective effect is affected by underlying medical conditions. Partial vaccination does not offer clinically meaningful protection against severe illness. Our study highlights the importance of continuing effort on a full course of vaccination.

## Data Availability

Some or all data, models, or code that support the findings of this study are available from the corresponding author upon reasonable request.

## Acknowledgements

We would like to thank all the heroic people for their active participation in ending this COVID-19 outbreak and for taking care of the patients.

